# Geographic Variation in Influenza Vaccination among US Nursing Home Residents: A National Study

**DOI:** 10.1101/2021.01.25.21250461

**Authors:** Joe B. B. Silva, Elliott Bosco, Melissa R. Riester, Kevin W. McConeghy, Patience Moyo, Robertus van Aalst, Barbara H. Bardenheier, Stefan Gravenstein, Rosa Baier, Matthew M. Loiacono, Ayman Chit, Andrew R. Zullo

## Abstract

**Objective:** Estimates of influenza vaccine use are not available at the county level for U.S. nursing home (NH) residents but are critically necessary to guide implementation of quality improvement programs aimed at increasing vaccination rates. Furthermore, estimates that account for differences in resident characteristics between counties are unavailable. We estimated risk-standardized vaccination rates among short- and long-stay NH residents by U.S. county and identified drivers of geographic variation.

**Methods:** We conducted a retrospective cohort study utilizing 100% of 2013-2015 fee-for-service Medicare claims, Minimum Data Set assessments, Certification and Survey Provider Enhanced Reports, and LTCFocUS. We separately evaluated short-stay (<100 days) and long-stay (≥100 days) residents aged ≥65 years old across the 2013-2014 and 2014-2015 influenza seasons. We estimated county-level risk-standardized vaccination rates (RSVRs) via hierarchical logistic regression adjusting for 32 resident-level covariates. We then used multivariable linear regression models to assess associations between county-level NHs predictors and RSVRs.

**Results:** The overall study cohort consisted of 2,817,217 residents in 14,658 NHs across 2,798 counties. Short-stay residents had lower RSVRs than long-stay residents (2013-2014: median [IQR], 69.6% [62.8-74.5] vs 84.0% [80.8-86.4]). Counties with the highest vaccination rates were concentrated in the Midwestern, Southern, and Northeast US. Several modifiable facility-level characteristics were associated with increased RSVRs, including higher registered nurse to total nurse ratio and higher total staffing for licensed practical nurses, speech language pathologists, and social workers. Characteristics associated with lower RSVRs included higher percentage of residents restrained, with a pressure ulcer, and NH-level hospitalizations per resident-year.

**Conclusions:** Substantial county-level variation in influenza vaccine use exists among short- and long-stay NH residents. Quality improvement interventions to improve vaccination rates can leverage these results to target NHs located in counties with lower risk-standardized vaccine use.

## INTRODUCTION

Older adults residing in nursing homes (NHs) are at high risk for respiratory infections, including influenza.^1-3^ Between 70% and 85% of influenza-related deaths occur in people aged 65 years and older.^4^ Influenza vaccination is considered a cost-effective method to prevent outbreaks in NHs. It decreases the risk and severity of infection and, in doing so, also decreases the risk of subsequent adverse outcomes such as hospitalization and mortality.^5^ However, current influenza vaccination rates among U.S. NH residents is well below the Healthy People 2020 goal of 90%.^6,7^ In order to implement effective and efficient quality improvement initiatives aimed at increasing influenza vaccine use in NHs, it is critical to first understand how influenza vaccination rates differ across the country and to identify areas with the lowest vaccine uptake. While influenza incidence rates are known to vary widely by U.S. county after adjusting for differences in residents between counties,^8^ the small-area geographic patterns of influenza vaccination among NH residents remain unknown. National data reporting geographic information for NH resident vaccination have provided national- or state-level estimates, but these mask variations in smaller areas, such as counties, and have other important limitations.^9,10^ Prior studies that have reported on person- or NH-level predictors of vaccination did not use risk-standardization when estimating influenza vaccination rates.^11-14^ Utilizing risk-standardization is critical as resident characteristics are known to vary across geographic units in the U.S. Without accounting for these differences across counties, it is extremely difficult to validly compare county-level vaccination rates or ascribe any observed differences to local policies and practices. Prior studies also have not analyzed short-stay (post-acute care) and long-stay (long-term care) residents as two distinct groups despite the two subpopulations’ markedly different care needs and profiles.^11-14^ Understanding vaccination rates for short- and long-stay residents separately is important, as drivers of vaccination and related interventions will likely differ for each of these populations. Indeed, many quality improvement interventions target each group separately.^15^Quantifying county-level vaccination rates is vital for local stakeholders responsible for making decisions to prevent and control infections in NHs. Absent such information, policymakers, clinicians, public health professionals, and organizations cannot allocate finite resources appropriately or focus quality improvement or other interventions on areas and specific NHs with the lowest vaccination rates. Failure to allocate resources and intervene effectively is likely to perpetuate any ongoing geographic inequalities in vaccine use among NH residents.

Accordingly, our objectives were to 1) estimate how risk-standardized influenza vaccination rates (RSVRs) among short- and long-stay NH residents varied across U.S. counties after adjusting for resident characteristics to ensure consistent and comparable estimates, and 2) identify potential drivers of observed small-area geographic variation. We hypothesized that wide variation in vaccination rates would exist across counties for both the short-stay and long-stay populations despite risk-standardization. We also hypothesized that modifiable NH-level predictors related to variety of staffing, direct care hours, and quality measures would be associated with increases in county-level RSVRs.

## METHODS

### Study Design and Data Source

This retrospective cohort study included fee-for-service Medicare beneficiaries who resided in U.S. NHs between January 1^st^, 2013 and December 31^st^, 2015. Data were derived from 100% samples of Medicare Parts A and B claims, Minimum Data Set (MDS) version 3.0 assessments, Certification and Survey Provider Enhanced Reports (CASPER), and Long-Term Care: Facts on Care in the U.S. (LTCFocUS).

Centers for Medicare & Medicaid Services- (CMS-) certified NHs are required to complete MDS resident assessments for all residents, to capture information including: demographics; clinical conditions; functional, psychological, and cognitive status; and services and treatments.^16^ Assessments must be performed at least quarterly for each resident, but are often performed more frequently; for example, with any change in condition.^16,17^ After linking assessment data to Medicare claims, we employed a previously-validated residential history file algorithm to track residents’ timing and location of health service utilization.^18^ We used CASPER and LTCFocUS data to capture NH-level information, including structural characteristics, staffing information, quality-of-care deficiencies, and aggregate resident characteristics. CASPER data are collected by state survey agencies during NH inspections. LTCFocUS is a product of the Shaping Long-Term Care in America Project at the Brown University Center for Gerontology & Healthcare Research and supported, in part, by the National Institute on Aging.^19,20^ The Brown University Institutional Review Board approved the study protocol.

### Study Population

We included beneficiaries aged 65 years or older on their index date (defined below) residing in free-standing (i.e., not hospital-based) NHs between January 1, 2013 and December 31, 2015 who had at least one MDS assessment. We also required that residents’ NH be identifiable, and the corresponding NH-level data be available in CASPER and LTCFocUS. Residents with less than six months of continuous enrollment in both Medicare Parts A and B or health maintenance organizations (i.e., Medicare Advantage) were excluded as were those with missing data on any person-level covariates necessary for risk-standardization (process described below in Statistical Analyses).

Residents were divided into four subcohorts according to their short-stay/long-stay status and influenza season of stay (2013-2014 and 2014-2015). First, residents were designated as either short-stay (SS) or long-stay (LS) based on length of residency in the NH. Short-stay residents were those with <100 days in the same NH and were assigned an index date of entry. Those designated as long-stay had ≥100 consecutive days with no more than 10 days outside of the NH and an index date assigned as day 100 of their stay. Stays were then assigned to seasons, where residents with at least one day of a NH stay from October 1, 2013 to March 31, 2014 were included in the 2013-2014 season subcohorts, while those with at least on day of a NH stay from October 1, 2014 to March 31, 2015 were included in the 2014-2015 season. Only residents’ first stay identified during a particular season was included, however residents could be represented in both seasons if they survived to the second and remained in the same NH.

### Resident Characteristics for Risk Adjustment

Resident characteristics were used in risk-standardization across counties. We selected 32 covariates encoding information about resident demographic information and comorbidities that were likely to be associated with receiving influenza vaccine.^2,21-23^ We obtained age, sex, and race from the Medicare enrollment file, and we derived all remaining demographic covariates and comorbidities from MDS assessments (Supplementary Table S1). For long-stay residents, covariates were derived from MDS assessments in the 100 days prior to their index date as well as all assessments after the index date during a given season. For short-stay residents, covariates were derived from all MDS assessments available from the time of admission to the NH to the time of discharge. Since risk-standardization requires complete resident-level data and the MDS Active Diagnoses Section I variables are not measured in every MDS assessment, this approach to MDS covariate ascertainment minimized missing data.

### Vaccination Status

Influenza vaccination status for vaccinations received October 1 to March 31 was ascertained from the immunization supplement on MDS resident assessments between October 1 and June 30. We emulated a previously-described algorithm to leverage responses from multiple MDS assessments and classify residents as vaccinated based two questions about influenza vaccination.^7,21^ These MDS questions included “Did the resident receive the influenza vaccine in this facility for this year’s influenza vaccination season?” and, subsequently, “If influenza vaccine was not received, state reason: (1) not in facility during this year’s flu season; (2) received outside of this facility; (3) not eligible; (4) offered and declined; (5) not offered; and (6) inability to obtain vaccine”.

### Nursing Home-Level Predictors for Examining Determinants of Vaccination

We obtained potential NH-level predictors of influenza vaccination from CASPER and LTCFocUS that have been previously described.^2,8,24,25^ We selected 28 predictors that included aggregate NH-level resident demographic predictors, potentially non-modifiable NH predictors, and potentially modifiable NH predictors (enumerated in Table 1). Aggregate NH-level resident demographic variables for all admissions included average age at index among the home’s residents, distribution of race and sex (percentage of White, Black, and Hispanic resident admissions, male vs. female admissions) within each NH, and average functional status within each NH, which was assessed via the 28-point Morris Activities of Daily Living (ADL) Scale.^26^ Nonmodifiable NH-level predictors that we included were number of beds, admissions per bed, occupancy rate, average daily census, resident primary payer (Medicare, Medicaid, or other), NH Acuity Index of residents,^27^ profit status of the NH, if the NH was part of a multi-facility corporation, and whether the NH was located in an urban vs. rural county.

**Table 1.** County-level NH Predictors for Short-Stay and Long-Stay Residents, Stratified by Influenza Season, N=2,798 counties.

| Predictor | 2013-2014 Season |  | 2014-2015 Season |  |
| --- | --- | --- | --- | --- |
|  | Short-Stay<br>(N=2,740) | Long-Stay<br>(N=2,786) | Short-Stay<br>(N=2,758) | Long-Stay<br>(N=2,781) |
| **Aggregate Resident Predictors** |  |  |  |  |
| Age at index date, years | 79.8 ± 2.8 | 83.9 ± 2.1 | 80.3 ± 2.8 | 83.8 ± 2.1 |
| % Female (all admissions) | 69.1 ± 7.2 | 68.6 ± 7.3 | 68.9 ± 7.5 | 68.5 ± 7.4 |
| % White (all admissions) | 86.1 ± 16.9 | 85.3 ± 17.2 | 86.1 ± 17.0 | 85.3 ± 17.3 |
| % Black (all admissions) | 7.8 ± 13.0 | 8.0 ± 13.2 | 7.9 ± 13.1 | 8.0 ± 13.2 |
| % Hispanic (all admissions) | 2.6 ± 9.5 | 2.6 ± 8.9 | 2.6 ± 9.4 | 2.6 ± 8.9 |
| ADL* | 16.1 ± 6.3 | 16.2 ± 6.5 | 16.1 ± 6.3 | 16.2 ± 6.4 |
| **Non-Modifiable NH Predictors** |  |  |  |  |
| Average Daily Census | 76.8 ± 30.6 | 75.6 ± 30.5 | 76.4 ± 30.5 | 75.5 ± 30.3 |
| Occupancy Rate | 80.5 ± 12.3 | 80.2 ± 12.7 | 80.5 ± 12.3 | 80.4 ± 12.5 |
| Total Beds | 97.0 ± 34.0 | 96.0 ± 34.0 | 97.0 ± 34.0 | 96.0 ± 34.0 |
| % Medicare | 13.1 ± 7.3 | 12.8 ± 7.2 | 12.8 ± 7.1 | 12.5 ± 6.9 |
| % Medicaid | 63.5 ± 13.1 | 63.4 ± 13.4 | 63.9 ± 12.9 | 63.9 ± 13.2 |
| % pay other | 23.4 ± 12.8 | 23.8 ± 13.0 | 23.4 ± 12.8 | 23.7 ± 12.9 |
| Admissions per bed | 1.6 ± 1.1 | 1.6 ± 0.8 | 1.5 ± 1.1 | 1.5 ± 0.8 |
| Average Acuity Index | 11.9 ± 1.1 | 12.0 ± 1.0 | 11.9 ± 1.2 | 12.0 ± 1.0 |
| % for Profit | 71.5 ± 34.4 | 70.8 ± 34.9 | 71.3 ± 34.6 | 70.7 ± 34.8 |
| % Multi-facility corporation | 58.4 ± 37.1 | 57.6 ± 37.0 | 59.0 ± 37.1 | 58.3 ± 37.1 |
| % Urban | 1,039 (38.0%) | 1,042 (37.4%) | 1,040 (37.7%) | 1,041 (37.4%) |
| **Modifiable NH Predictors** |  |  |  |  |
| CNA to nurse ratio | 2.1 ± 0.5 | 2.0 ± 0.5 | 2.1 ± 0.5 | 2.0 ± 0.5 |
| RN to total nurse ratio | 0.3 ± 0.2 | 0.3 ± 0.2 | 0.3 ± 0.2 | 0.3 ± 0.2 |
| Total nursing hours per resident day | 3.9 ± 1.8 | 3.8 ± 1.4 | 3.8 ± 1.8 | 3.8 ± 1.3 |
| Hospitalizations per resident year | 1.0 ± 0.5 | 1.0 ± 0.5 | 1.0 ± 0.5 | 1.0 ± 0.5 |
| CNA hours per resident day | 2.3 ± 0.5 | 2.3 ± 0.5 | 2.3 ± 0.5 | 2.3 ± 0.5 |
| LPN hours per resident day | 0.8 ± 0.3 | 0.8 ± 0.3 | 0.8 ± 0.3 | 0.8 ± 0.3 |
| Total physician extender FTEs/100 beds | 0.1 ± 0.2 | 0.1 ± 0.2 | 0.1 ± 0.2 | 0.1 ± .02 |
| SLP total staff FTE | 335.0 ± 748.4 | 318.0 ± 667.9 | 343.8 ± 760.0 | 323.1 ± 669.3 |
| Social worker on staff/100 beds | 1.0 ± 0.8 | 1.0 ± 0.9 | 1.0 ± 0.8 | 1.0 ± .09 |
| Pressure ulcers | 5.7 ± 3.1 | 5.8 ± 3.2 | 5.7 ± 3.1 | 5.7 ± 3.2 |
| Restraints | 2.6 ± 5.0 | 2.1 ± 4.1 | 2.7 ± 5.2 | 2.1 ± 4.0 |

**Note:** The data are presented as either mean (standard deviation) or count (%).
**Abbreviations:** NH, nursing home; SD, standard deviation; CNA, certified nursing assistant; RN, registered nurse; LPN, licensed practical nurse; FTE, full-time equivalent; ADL, activities of daily living.
\*Functional status was assessed by calculating the 28-point Morris Activities of Daily Living (ADL) Scale.<sup>26</sup>

Modifiable NH-level predictors were those characteristics we identified as potential intervention targets and included several aspects of NH staffing, such as on registered nurses (RNs) and certified nursing assistants (CNAs) employed. We defined an array of NH-level measures of workforce variety, such as the staffing ratio of RNs to total nurses (RNs, CNAs and licensed practical nurses, or LPNs), the ratio of CNAs to total nurses, total social worker full-time equivalents (FTEs) per 100 beds, total physician extender full-time equivalents (FTE) per 100 beds, and speech language pathologist (SLP) total FTEs. We captured direct care hours via the total nursing hours/resident/day, CNA hours/resident/day, and LPN hours/resident/day. Finally, we included variables related to quality measures, such as the percentage of residents who were restrained, the percentage of residents who had a pressure ulcer, and the NH’s rate of hospitalizations per resident-year.

### County-Level Predictors for Examining Determinants of Vaccination

We selected the county as the geographic unit of analysis because beneficiaries typically choose NHs at the county level and it represents the smallest geographic unit with policy implications. It is also likely that successful quality improvement interventions aiming to improve vaccination rates may need to be implemented at the county level to maximize effectiveness. NHs and residents were assigned to counties based on the zip code of the NH, which were then aggregated up to the level of Federal Information Processing Standards county codes.

### Statistical Analyses

We calculated crude vaccination rates and RSVRs for each county. Risk-standardization using the resident-level covariates was necessary to adjust for well-known resident-level differences between counties. Adapting from CMS methodology for risk-standardization, county-level RSVRs were calculated by multiplying the ratio of predicted to expected influenza vaccinations by the crude influenza vaccinations for each subcohort.^8,24,28^ Hierarchical logistic regression models adjusted for 32 resident-level covariates were fit for each of the four subcohorts in the study (short-stay 2013-2014, long-stay 2013-2014, short-stay 2014-2015, long-stay 2014-2015) to estimate predicted and expected influenza vaccinations with a county-specific intercept and an *average* county-specific intercept based on all counties within in each subcohort.

Geospatial analyses were then conducted to explore the geographic patterns of crude vaccination rates and RSVRs. Choropleth maps were generated for each of the four subcohorts to plot crude vaccination rates and RSVR quintiles. Analyses were separated by subcohort, therefore the cut-points for quintiles represented on maps were not the same across seasons or short-stay and long-stay populations.

Finally, multivariable linear regression models using ordinary least squares were used to evaluate the associations between county-level RSVRs and 28 county-level predictors. The models provided percent differences in vaccination rates and 95% confidence intervals for each of the four subcohorts in the study. For continuous predictors, counties were grouped into tertiles with inferences made based on the percent differences in RSVRs among counties grouped in the highest tertile of a specific predictor compared counties in the lowest tertile. Predictors were classified as associated with increases or decreases in RSVRs if the direction of association was present in at least three of the four subcohorts.

### Stability Analyses

We evaluated alternative approaches to determine the impact of including counties with few NH residents on the stability of the main county-level RSVR estimates. In the first stability analysis, we excluded all counties with fewer than 5 total residents. In the second stability analysis, we excluded counties with fewer than 7. This was analogous to making a trade-off between precision and generalizability since estimates from counties with more NH residents were expected to be more precise, but potentially less generalizable to the target population of all counties with NHs across the U.S. In our multivariable linear regression models, we also evaluated the impact of using Huber-White standard errors on the inferences drawn.

### Software

We conducted analyses using SAS Enterprise Guide 8.1 (SAS Institute, Cary, North Carolina), R version 3.6.1 (R Foundation for Statistical Computing, Vienna, Austria), and ArcMap 10.8 (ESRI, Redlands, California) software.

## RESULTS

### Study Cohort and Vaccination Rates

The overall study cohort consisted of 2,817,217 residents in 14,658 NHs across 2,798 counties. During the 2013-2014 season, there were 524,739 short-stay residents in 14,059 NHs across 2,740 counties, and 851,869 long-stay residents in 14,494 NHs across 2,789 counties (Table 1; Supplementary Figure S1). The subcohorts for the 2014-2015 season consisted of 689,061 short-stay residents in 14,150 NHs across 2,758 counties, and 881,264 long-stay residents in 14,463 NHs across 2,781 counties (Table 1; Supplementary Figure S1).

The median county-level crude vaccination rate among short-stay residents during the 2013-2014 season was 73.2% (interquartile range, [IQR] 61.3-81.8) and 72.7% (IQR, 60.9-81.2) during the 2014-2015 season. For long-stay residents, the median county-level crude vaccination rate was 86.8% (IQR, 82.0-90.5) during the 2013-2014 season and 85.7% (IQR, 81.0-93.0) during the 2014-2015 season (Table 2). After risk adjustment, the median county-level RSVR among short-stay residents in 2013-2014 was 69.6% (IQR, 62.8-74.5) and 69.1% (IQR, 62.0-74.1) in 2014-2015. Across long-stay residents, the median county-level RSVR in 2013-2014 was 84.0% (IQR, 80.8-86.4) and 83.1% (IQR, 79.7-85.7) in 2014-2015 (Table 2).

**Table 2.** Crude and Risk-Standardized County-Level Influenza Vaccination Rates of Short-Stay and Long-Stay Residents, Stratified by Season, N=2,798 counties.

|  | **2013-2014 Season** |  | **2014-2015 Season** |  |
| --- | --- | --- | --- | --- |
|  | **Short-Stay**<br>**(N=2,740)** | **Long-Stay**<br>**(N=2,786)** | **Short-Stay**<br>**(N=2,758)** | **Long-Stay**<br>**(N=2,781)** |
| Crude Assessed and Provided* | 92.0 (84.6, 97.2) | 97.6 (95.7, 99.1) | 91.4 (83.5, 96.2) | 97.3 (95.2, 98.9) |
| Crude Declined | 13.9 (7.0, 22.2) | 9.1 (6.3, 12.8) | 14.0 (7.7, 22.1) | 9.6 (6.6, 13.5) |
| Crude Contraindicated | 0 (0, 1.3) | 0.8 (0, 1.5) | 0.0 (0, 1.3) | 0.8 (0, 1.5) |
| Crude Did Not Offer | 0 (0, 3.1) | 0 (0, 1.0) | 0.3 (0, 3.1) | 0 (0, 1.1) |
| Crude Vaccinated | 73.2 (61.3, 81.8) | 86.8 (82.0, 90.5) | 72.7 (60.9, 81.2) | 85.7 (81.0, 93.0) |
| Risk-Standardized Vaccinated | 69.6 (62.8, 74.5) | 84.0 (80.8, 86.4) | 69.1 (62.0, 74.1) | 83.1 (79.7, 85.7) |

**Note:** The data are presented as median (interquartile range).
\*Assessed and Provided is an additive measure that takes into account those who were vaccinated, declined vaccination, or who had a contraindication to receiving the vaccine.

### Geospatial Analysis

Wide geographic variation was observed for both crude and risk-standardized vaccination rates across both the short- and long-stay populations and both influenza seasons (Figure 1, Figure 2). While observed patterns were consistent between crude and risk-standardized choropleth maps for subcohorts, 65.80% of counties (n=1,841) experienced one change in vaccination rate quintile (to an adjacent quintile) after risk-standardization in at least one subcohort. A change of at least two vaccination rate quintiles (to a non-adjacent quintile) for a given subcohort was observed in 112 counties, which were found primarily in a band stretching from northern Texas through counties in Oklahoma, Kansas, South Dakota, and North Dakota. Two counties, Jefferson county, Mississippi and Gadsden county, Florida, shifted three vaccination rate quintiles through risk-standardization for the long-stay population in the 2014-2015 influenza season.

**Figure 1.**
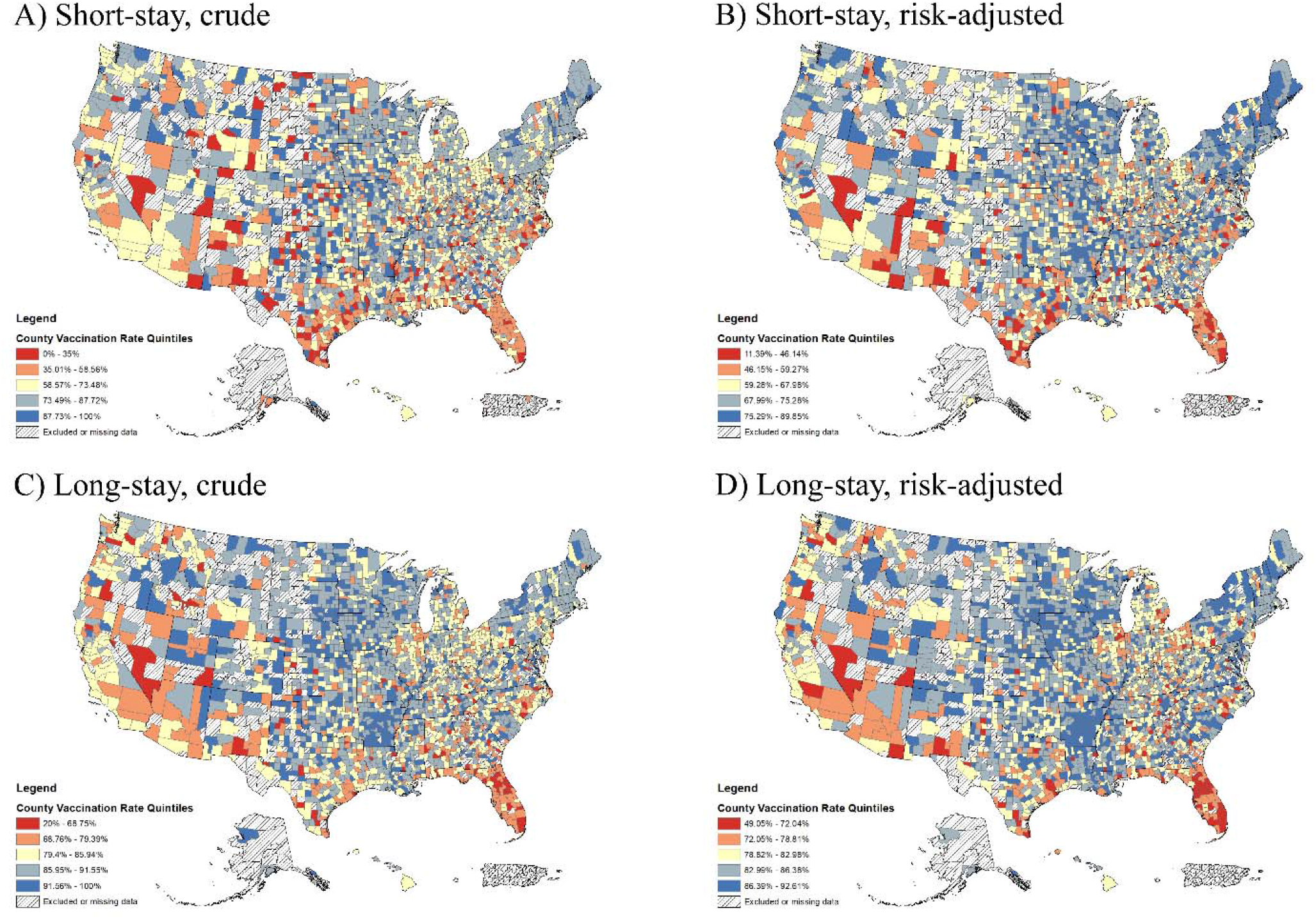
Crude and Risk-Standardized County-Level Vaccination Rates among Short-Stay and Long-Stay Nursing Home Residents during 2013-2014 Influenza Season, N=2,788 counties.

**Figure 2.**
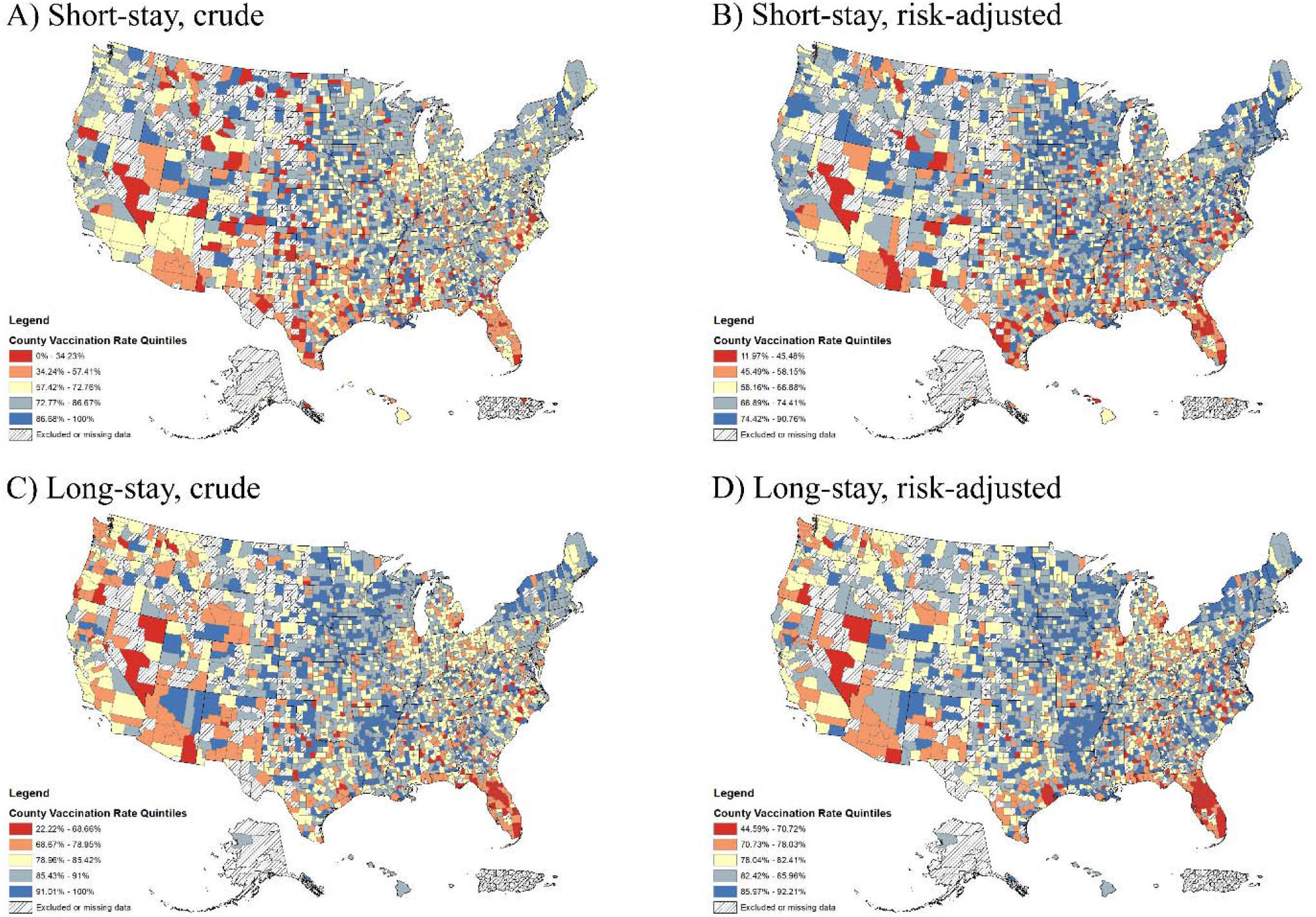
Crude and Risk-Standardized County-Level Vaccination Rates among Short-Stay and Long-Stay Nursing Home Residents during 2014-2015 Influenza Season, N=2,783 counties.

Counties with the highest RSVRs were located predominately in the Midwest (central Iowa, southern Minnesota, western Wisconsin), the South (central Arkansas, Louisiana, Georgia, and Virginia), and parts of the Northeast (counties in Finger Lakes and Central regions of New York) (Figure 1, Panels B and D; Figure 2, Panels B and D). Counties with the lowest RSVRs were particularly concentrated in counties in the South, including a band stretching from Houston, Texas along the Gulf of Mexico through much of Florida, and another along the Inner Coastal Plain of North Carolina (Figure 1, Panels B and D; Figure 2, Panels B and D). Counties with low vaccination rates were also located throughout Tennessee, Kentucky, Georgia, and Virginia as well as metro areas around Las Vegas, Nevada; Phoenix, Arizona; Los Angeles, California; and Chicago, Illinois.

The observed patterns of vaccine use were consistent across seasons for short- and long-stay populations, but county-specific RSVRs varied from 2013-2014 to 2014-2015 (Figure 1, Figure 2). Among the long-stay population, three counties, Valencia, New Mexico; Pittsylvania, Virginia; and Perquimans, North Carolina, had a difference of greater than 20 percentage points in RSVRs between seasons (Figure 1, Panel D; Figure 2, Panel D). In the short-stay population, differences of 20 percentage points or more occurred in 135 counties distributed across parts of Georgia, North Carolina, Mississippi, Illinois, Missouri, Oklahoma, and Texas (Figure 1, Panel B; Figure 2, Panel B).

### Multivariable Analyses

Among aggregate resident-level predictors, higher average age at index, higher proportion of Black resident admissions, and higher percentage of female resident admissions were associated with increases in RSVRs among all subcohorts (Table 3, Supplementary Table S2). Higher percentage of Hispanic resident admissions was associated with decreased RSVRs in all subcohorts (SS13-14: -2.07%, SS14-15: -1.95%, LS13-14: -1.41%, LS14-15: -1.36%). Higher percentage of White resident admissions was associated with increased RSVRs in short-stay (SS13-14: 1.86%, SS14-15: 2.96%) but decreases in long-stay (SS13-14: -0.25%, SS14-15: - 0.13%), while higher average functional status was associated with increases in long-stay but decreases in short-stay.

**Table 3.**
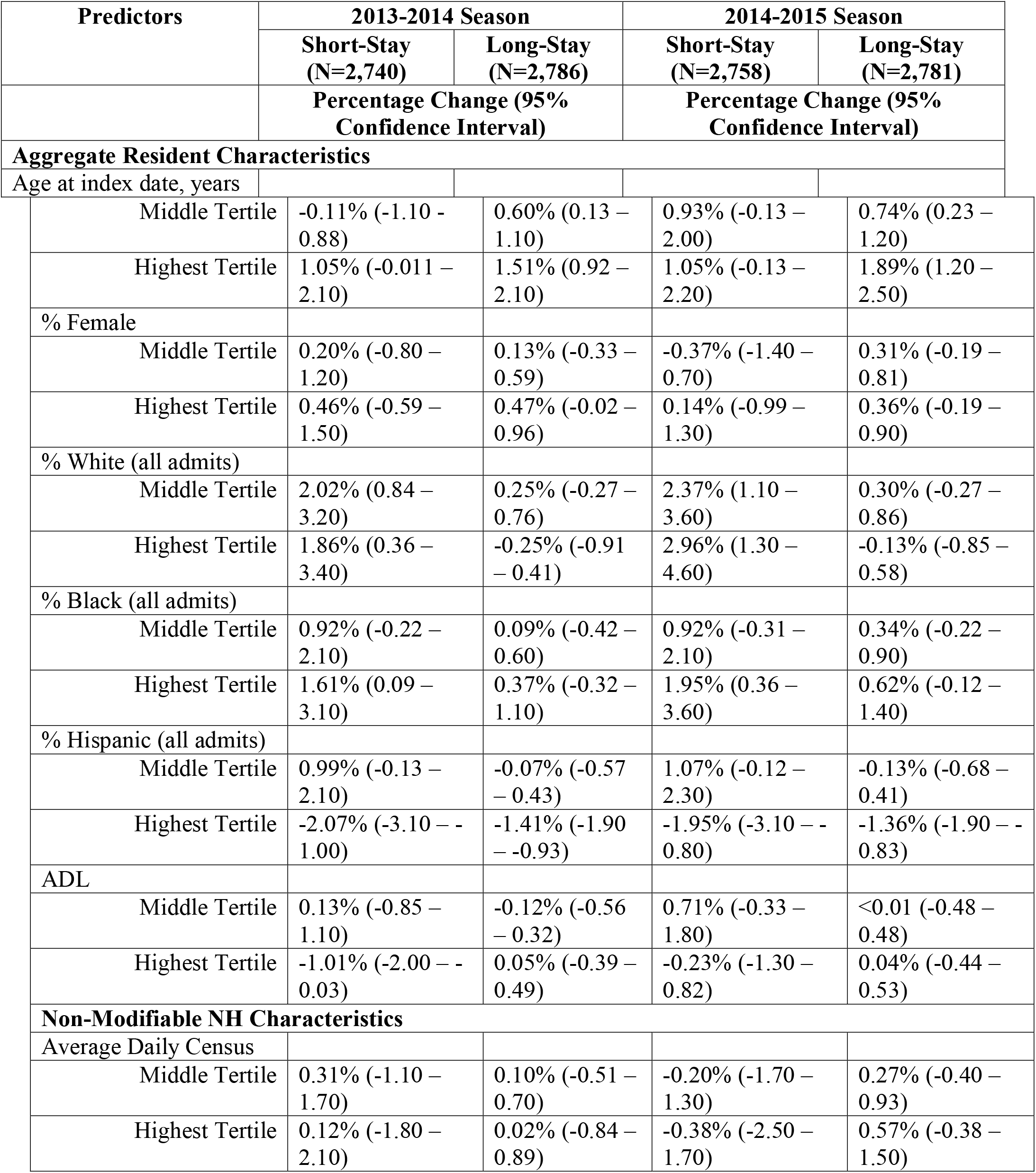
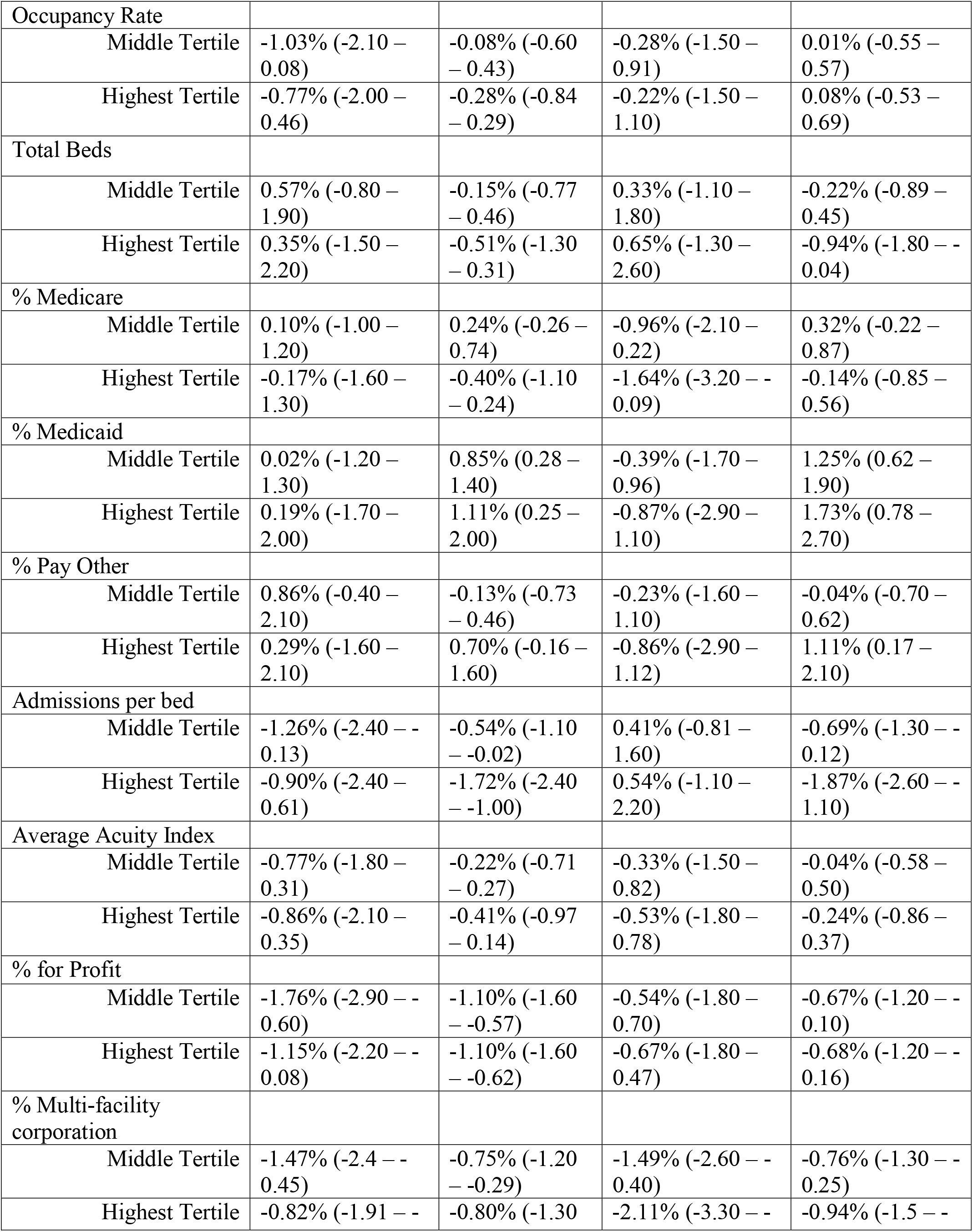
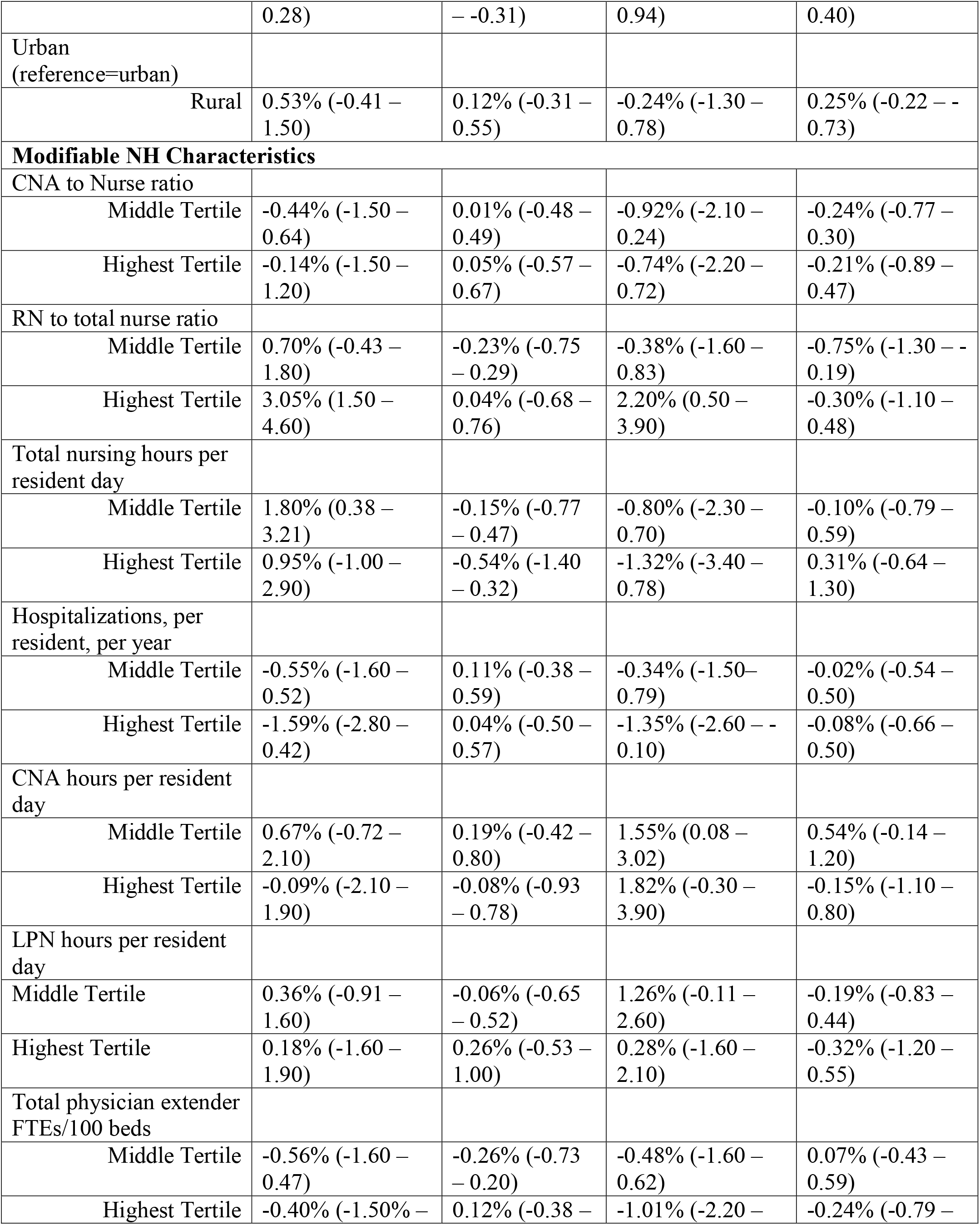
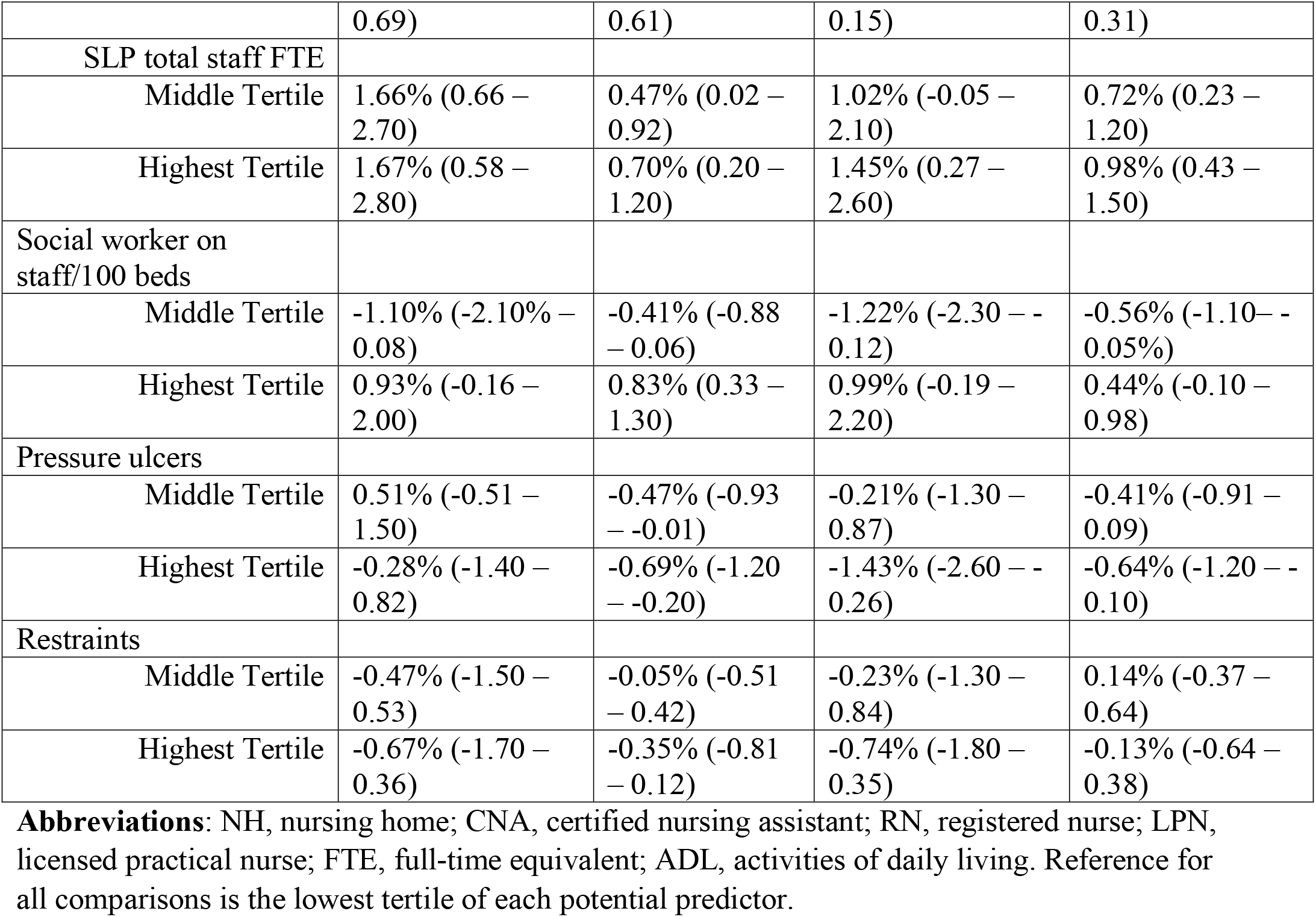
Multivariable Linear Regression Analyses to Identify Factors Associated with County-level Risk-Standardized Influenza Vaccination Rates Among Short-Stay and Long-Stay NH Residents, Stratified by Season, N=2,798 counties.

Non-modifiable NH-level predictors that were associated with increases in RSVRs included higher percentage of Medicaid residents (SS13-14: 0.19%, SS14-15: -0.87%, LS13-14: 1.11%, LS14-15: 1.73%), higher percentage of other non-Medicare non-Medicaid payer residents, higher average daily census counts, and counties classified as rural settings. Non-modifiable NH-level predictors associated with decreases in RSVRs included higher percentage of NHs part of multi-facility corporations within the county sample (SS13-14: -0.82%, SS14-15: -2.11%, LS13-14: -0.80%, LS14-15: -0.94%), higher occupancy rates, higher percentage of Medicare residents, higher admissions per bed, higher average acuity index, and higher percentage of for-profit NHs within the county. Higher total bed count was associated with increased RSVRs in the short-stay population but decreased RSVRs in the long-stay population.

Modifiable NH-level predictors that were associated with higher RSVRs included higher RN to total nurse ratio (SS13-14: 3.05%, SS14-15: 2.20%, LS13-14: 0.04%, LS14-15: -0.30%), higher total LPN hours, higher total SLP FTEs, and higher total social worker FTEs/100 beds. Decreases in RSVRs were found to be associated with higher average rate of hospitalizations per resident-year (SS13-14: -1.59%, SS14-15: -1.35%, LS13-14: 0.04%, LS14-15: -0.08%), higher percentage of residents who had a pressure ulcer, higher percentage of residents who were restrained, higher total CNA hours, higher CNA to total nurse ratio, and higher total physician extender FTE/100 beds. Higher total nursing hours hours/resident/day was associated with increased RSVRs the short-stay 2013-2014 and long-stay 2014-2015 subcohorts but decreases in the long-stay 2013-2014 and short-stay 2014-2015 subcohorts.

### Stability Analysis

The stability analysis (Supplementary Table S3) produced results that were consistent with the main findings. While excluded counties were predominately excluded in analyses of the short-stay subcohorts, there was no significant difference in the median RSVR or geographic variation across counties. Our findings were also consistent when using Huber-White standard errors for our multivariable linear regression models.

## DISCUSSION

In this large national retrospective cohort study of NH residents from 2013 to 2015, we found notable geographic variation in vaccination rates for both the short- and long-stay populations. The foundational evidence we provide can inform strategic efforts at the state- and county-levels to improve the health outcomes of NH residents by increasing influenza vaccination. Our findings also reveal that crude estimates of vaccination rates were optimistic, and that accounting for differences in resident-level characteristics across counties changed inferences about which counties might be the best targets for NH quality improvement interventions.

The geospatial variation in vaccination rates that we documented among older adults in NHs is similar to that observed for the general population.^29^ It is also generally consistent with previously documented state-level variation in vaccination among NH residents.^9,10^ Our results related to the drivers of vaccination in NHs concord with prior work establishing that vaccination rates are higher in NHs with better staffing.^24,30,31^ We found that increased RN to total nurse ratio, LPN hours per resident day, social worker FTEs per 100 beds, and total SLP FTEs at the county-level were associated with increased RSVRs. Our findings also agree with other studies of important drivers of the associations between vaccination rates and non-modifiable NH-level predictors. NHs in rural counties, NHs operated by the government or non-profit organizations, NHs not affiliated with multi-facility corporations, and NHs with fewer Medicare residents all had higher vaccination rates.^13,14^

While prior studies have described state-level NH resident vaccination rates, our findings reveal new patterns of heterogeneity in vaccination rates at the county-level. High- and low-vaccinating counties were identified in most states such that, for example, subregions of high-vaccinating counties exist in low-vaccinating states. Examples of such subregions are those surrounding Tallahassee, Florida, and counties between San Antonio and Austin, Texas. Among states in the middle distribution of state-level variation in vaccination among NH residents, more heterogeneity across counties was observed. South Carolina, ranked in the middle quintile for resident vaccination,^9,10^ contains counties with high RSVRs in the westernmost regions of the state, and counties with lower RSVRs among coastal communities. More studies are needed to understand the determinants increased heterogeneity among vaccination rates at the county-level within states.

### Limitations

The findings of this study must be interpreted in light of several limitations.

First, while observed patterns of NH geospatial vaccine use were comparable between seasons, it is unknown if these patterns are generalizable to the 2015-2016 season and after. Resident influenza vaccine use may have increased over time, but it is unclear if use has continued to increase at the same rate across states and counties.^32^ Since analyses were limited to Medicare beneficiaries aged 65 years or older, the results may also not generalize to younger individuals with disabilities who reside in NHs. However, individuals aged 65 years or older represent the majority of residents.^33^

Second, some counties have a higher number of total NH residents, which influences the precision of risk-standardized vaccination rates across counties. Our two stability analyses, which restricted to counties with a minimum of five or seven NH residents, respectively, provided evidence that this may not be a major concern because inferences were generally unaffected.

Third, we identified several county-level predictors that may drive the observed patterns of intra-state variation, however, other potential drivers such as hospital-NH relationships, county-level incidence of influenza, and county-level NH-staff vaccination rates may also influence local vaccine use.^8,24^ Staff factors are likely to be particularly important. Staff vaccination can not only lessen severity of illness and prevent spread through herd-immunity, but also prevent absenteeism in the presence of an outbreak.^5,34^ Current CMS regulations include requirements for offering residents vaccination, but state public health laws also exist and may consist of assessment requirements, administrative requirements for offering vaccination, and administrative requirements to ensure vaccination of both NH residents and staff.^35,36^ Arkansas, a state with many high performing counties, and Florida, a state with many low performing counties, had similar influenza vaccination laws for residents, but Florida had no existing state law for NH staff vaccination.^36^

Finally, the associations we estimate between county-level predictors and vaccination are not estimates of causal effects and should not be interpreted as such. Additional work would be necessary for causal inference.

## Conclusions and Implications

Our results can be used to deploy quality improvement and other interventions to improve influenza vaccination rates in NHs at the county level. Current interventions that might be deployed using our findings include The Immunization Champions, Advocates and Mentors Program (ICAMP), which is a multidisciplinary program for healthcare professionals who are committed to improving vaccination rates.^37^ Others certainly also exist. In order to disseminate ICAMP or other quality improvement interventions, qualitative research exploring care processes and practices in both the highest RSVR and lowest RSVR counties is one of several ideal next steps towards further uncovering why differences exist between high and low performing counties. While some successful local interventions to improve NH resident vaccination have been previously described,^38^ an additional understanding of which practices work well in which settings can likely spur progress toward further reducing the disparities that exist between counties. Finally, our findings may also be relevant for guiding the distribution and uptake of COVID-19 and other vaccines if similar geographic patterns exist, though future work should be conducted.

In summary, wide county-level geographic variation in influenza vaccine use among long-stay and short-stay NH residents was observed in this study. In combination with information about the geospatial variation in influenza in NHs, this information about the geospatial variation in vaccine use should be used by local public health authorities and clinicians to target interventions to improve vaccine use to counties with both high rates of infection and low rates of vaccine use in their NHs. The results can also be used to identify areas that are in need of other interventions to reduce infections, such as better infection preventionist staffing models. Additional research is warranted to further elucidate the sources of geographic variation in vaccine use and support improved health outcomes of NH residents.

## Data Availability

The data were used under Data Use Agreement with the Centers for Medicare and Medicaid Services and are not available for sharing.

