## Supplemental Tables and Figures for "Geographic Variation in Influenza Vaccination among US Nursing Home Residents: A National Study"

**Supplementary Table S1. Covariates Included in Risk-adjustment.**

**Resident-level Characteristics**

Age at index

Female

Race/ethnicity (non-Hispanic White, non-Hispanic Black, Hispanic, Other)

BMI

**MDS comorbidities active in last 7 days**

Cancer

Atrial fibrillation or other dysrhythmias

Coronary artery disease

Deep vein thrombosis, pulmonary embolism,
 or pulmonary thromboembolism

Cerebrovascular accident, transient ischemic attack, or stroke

Heart failure

Hypertension

Gastroesophageal reflux disease or ulcer

Diabetes mellitus

Alzheimer’s disease

Non-Alzheimer’s Dementia

Parkinson’s Disease

Asthma, Chronic Obstructive Pulmonary Disease,
Chronic Lung Disease

Respiratory Failure

Pneumonia

Tuberculosis

Septicemia

Fever

Anemia

Arthritis

Crohn’s/Ulcerative Colitis

Multiple Sclerosis

Tube Feeding

Depression

Manic Depression

PTSD

Schizophrenia

Psychotic Disorder

**Abbreviations:** BMI, body mass index; MDS, minimum data set; PTSD, post-traumatic stress disorder.

**Supplementary Figure S1. Consort Diagram of Study Cohort, Stratified by Influenza Season, N =2,817,217.**

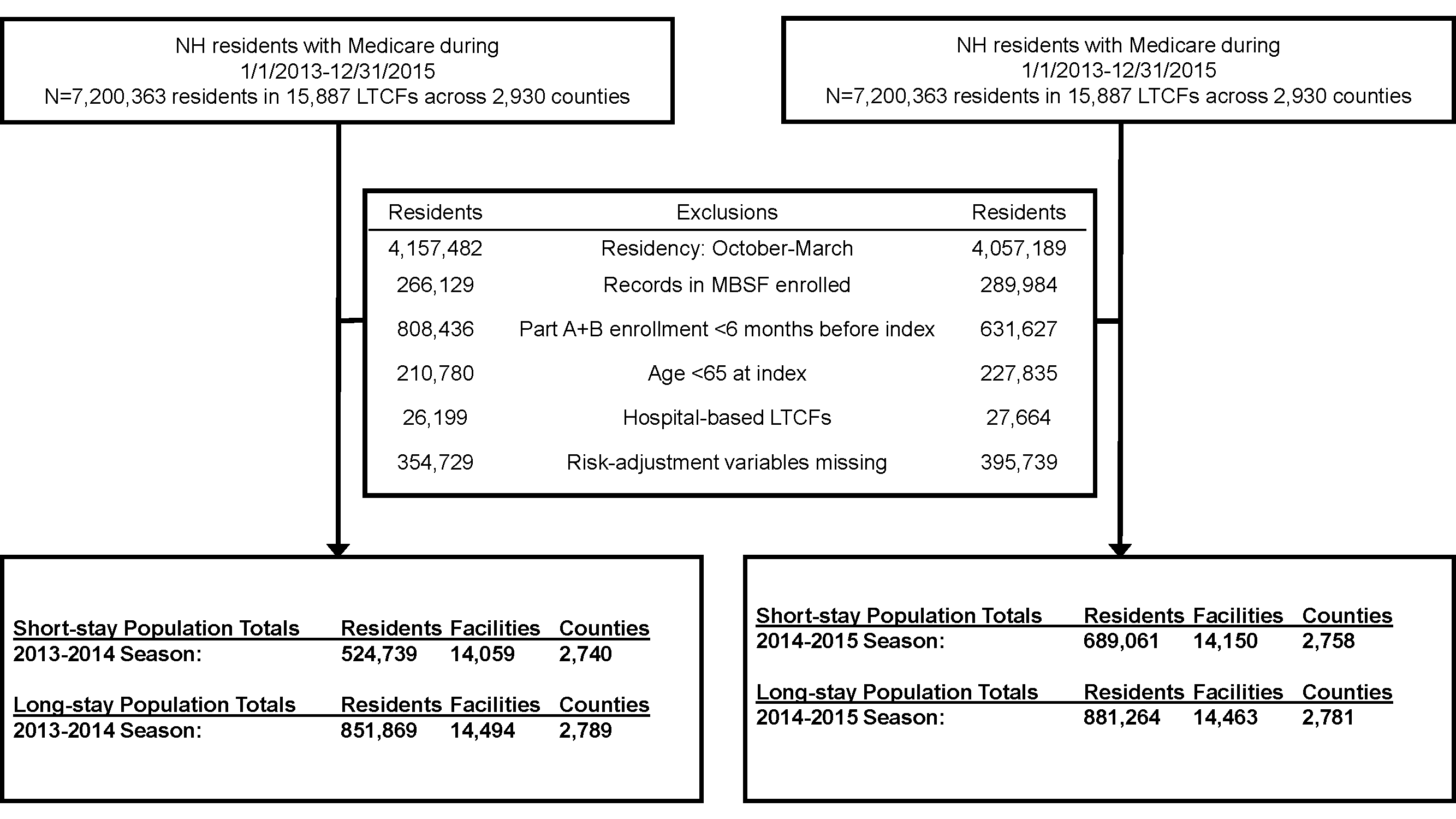

**Abbreviations:** NH, nursing home; MBSF, master beneficiary summary file.

**Supplementary Table S2. Multivariable Linear Regression Analyses to Identify Factors Associated with County-level Risk-Standardized Influenza Vaccination Rates Among Short-Stay and Long-Stay NH Residents, Stratified by Season, N=2,798 counties.**

| **Predictor** | **2013-2014 Season** | | **2014-2015 Season** | |
| --- | --- | --- | --- | --- |
|  | **Short-Stay**  **(N=2,740)** | **Long-Stay**  **(N=2,786)** | **Short-Stay**  **(N=2,758)** | **Long-Stay**  **(N=2,781)** |
|  | **Coefficient (95% Confidence Interval)** | | **Coefficient (95% Confidence Interval)** | |
| **Aggregate Resident Characteristics** | | | | |
| Age at index date, years |  |  |  |  |
| Middle Tertile | -0.0011 (-0.011 - 0.0088) | 0.00602* (0.0013 – 0.011) | 0.00933 (-0.0013 – 0.020) | 0.00738* (0.0023 – 0.012) |
| Highest Tertile | 0.01053 (-0.00011 – 0.021) | 0.01512* (0.0092 – 0.021) | 0.01045 (-0.0013 – 0.022) | 0.01887* (0.012 – 0.025) |
| % Female |  |  |  |  |
| Middle Tertile | 0.00198 (-0.0080 – 0.012) | 0.00128 (-0.0033 – 0.0059) | -0.0037 (-0.014 – 0.0070) | 0.00311 (-0.0019 – 0.0081) |
| Highest Tertile | 0.00456 (-0.0059 – 0.015) | 0.00471 (-0.00019 – 0.0096) | 0.00137 (-0.0099 – 0.013) | 0.00356 (-0.0019 – 0.0090) |
| % White (all admits) |  |  |  |  |
| Middle Tertile | 0.02017* (0.084 – 0.032) | 0.00246 (-0.0027 – 0.0076) | 0.02374* (0.011 – 0.036) | 0.00295 (-0.0027 – 0.0086) |
| Highest Tertile | 0.01861* (0.0036 – 0.034) | -0.0025 (-0.0091 – 0.0041) | 0.02956* (0.013 – 0.046) | -0.0013 (-0.0085 – 0.0058) |
| % Black (all admits) |  |  |  |  |
| Middle Tertile | 0.00922 (-0.0022 – 0.021) | 0.00089 (-0.0042 – 0.0060) | 0.00917 (-0.0031 – 0.021) | 0.00339 (-0.0022 – 0.0090) |
| Highest Tertile | 0.0161* (0.00088 – 0.031) | 0.00368 (-0.0032 – 0.011) | 0.01951* (0.0036 – 0.036) | 0.00624 (-0.0012 – 0.014) |
| % Hispanic (all admits) |  |  |  |  |
| Middle Tertile | 0.00988 (-0.0013 – 0.021) | -0.0007 (-0.0057 – 0.0043) | 0.01074 (-0.0012 – 0.023) | -0.0013 (-0.0068 – 0.0041) |
| Highest Tertile | -0.0207* (-0.031 – -0.010) | -0.0141* (-0.019 – -0.0093) | -0.0195* (-0.031 – -0.0080) | -0.0136* (-0.019 – -0.0083) |
| ADL |  |  |  |  |
| Middle Tertile | 0.00131 (-0.0085 – 0.011) | -0.0012 (-0.0056 – 0.0032) | 0.00711 (-0.0033 – 0.018) | -0.000028 (-0.0048 – 0.0048) |
| Highest Tertile | -0.0101* (-0.020 – -0.00025) | 0.00051 (-0.0039 – 0.0049) | -0.0023 (-0.013 – 0.0082) | 0.00044 (-0.0044 – 0.0053) |
| **Non-Modifiable NH Characteristics** | | | | |
| Average Daily Census |  |  |  |  |
| Middle Tertile | 0.00307 (-0.011 – 0.017) | 0.001 (-0.0051 – 0.0070) | -0.002 (-0.017 – 0.013) | 0.00267 (-0.0040 – 0.0093) |
| Highest Tertile | 0.00121 (-0.018 – 0.021) | 0.00021 (-0.0084 – 0.0089) | -0.0038 (-0.025 – 0.017) | 0.00567 (-0.0038 – 0.015) |
| Occupancy Rate |  |  |  |  |
| Middle Tertile | -0.0103 (-0.021 – 0.00084) | -0.0008 (-0.0060 – 0.0043) | -0.0028 (-0.015 – 0.0091) | 0.00011 (-0.0055 – 0.0057) |
| Highest Tertile | -0.0077 (-0.020 – 0.0046) | -0.0028 (-0.0084 – 0.0029) | -0.0022 (-0.015 – 0.011) | 0.00078 (-0.0053 – 0.0069) |
| Total Beds |  |  |  |  |
| Middle Tertile | 0.00569 (-0.0080 – 0.019) | -0.0015 (-0.0077 – 0.0046) | 0.00333 (-0.011 – 0.018) | -0.0022 (-0.0089 – 0.0045) |
| Highest Tertile | 0.00354 (-0.015 – 0.022) | -0.0051 (-0.013 – 0.0031) | 0.00645 (-0.013 – 0.026) | -0.0094* (-0.018 – -0.00043) |
| % Medicare |  |  |  |  |
| Middle Tertile | 0.00096 (-0.010 – 0.012) | 0.0024 (-0.0026 – 0.0074) | -0.0096 (-0.021 – 0.0022) | 0.00323 (-0.0022 – 0.0087) |
| Highest Tertile | -0.0017 (-0.016 – 0.013) | -0.004 (-0.011 – 0.0024) | -0.0164* (-0.032 – -0.0009) | -0.0014 (-0.0085 – 0.0056) |
| % Medicaid |  |  |  |  |
| Middle Tertile | 0.00019 (-0.012 – 0.013) | 0.00845* (0.0028 – 0.014) | -0.0039 (-0.017 – 0.0096) | 0.01254* (0.0062 – 0.019) |
| Highest Tertile | 0.00189 (-0.017 – 0.020) | 0.01109* (0.0025 – 0.020) | -0.0087 (-0.029 – 0.011) | 0.01727* (0.0078 – 0.027) |
| % Pay Other |  |  |  |  |
| Middle Tertile | 0.00863 (-0.0040 – 0.021) | -0.0013 (-0.0073 – 0.0046) | -0.0023 (-0.016 – 0.011) | -0.0004 (-0.0070 – 0.0062) |
| Highest Tertile | 0.00286 (-0.016 – 0.021) | 0.00701 (-0.0016 – 0.016) | -0.0086 (-0.029 – 0.012) | 0.01113* (0.0017 – 0.021) |
| Admissions per bed |  |  |  |  |
| Middle Tertile | -0.0126* (-0.024 – -0.0013) | -0.0054* (-0.011 – -0.00022) | 0.00407 (-0.0081 – 0.016) | -0.0069* (-0.013 – -0.0012) |
| Highest Tertile | -0.009 (-0.024 – 0.0061) | -0.0172* (-0.024 – -0.010) | 0.0054 (-0.011 – 0.022) | -0.0187* (-0.026 – -0.011) |
| Average Acuity Index |  |  |  |  |
| Middle Tertile | -0.0077 (-0.018 – 0.0031) | -0.0022 (-0.0071 – 0.0027) | -0.0033 (-0.015 – 0.0082) | -0.0004 (-0.0058 – 0.0050) |
| Highest Tertile | -0.0086 (-0.021 – 0.0035) | -0.0041 (-0.0097 – 0.0014) | -0.0053 (-0.018 – 0.0078) | -0.0024 (-0.0086 – 0.0037) |
| % for Profit |  |  |  |  |
| Middle Tertile | -0.0176* (-0.029 – -0.0060) | -0.011* (-0.016 – -0.0057) | -0.0054 (-0.018 – 0.0070) | -0.0067* (-0.012 – -0.0010) |
| Highest Tertile | -0.0115* (-0.022 – -0.00084) | -0.011* (-0.016 – -0.0062) | -0.0067 (-0.018 – 0.0047) | -0.0068* (-0.012 – -0.0016) |
| % Multi-facility corporation |  |  |  |  |
| Middle Tertile | -0.0147* (-0.024 – -0.0045) | -0.0075* (-0.012 – -0.0029) | -0.0149* (-0.026 – -0.0040) | -0.0076* (-0.013 – -0.0025) |
| Highest Tertile | -0.0082* (-0.019 – 0.0028) | -0.008* (-0.013 – -0.0031) | -0.0211* (-0.033 – -0.0094) | -0.0094* (-0.015 – -0.0040) |
| Urban (reference=urban) |  |  |  |  |
| Rural | 0.00533 (-0.0041 – 0.015) | 0.00119 (-0.0031 – 0.0055) | -0.0024 (-0.013 – 0.0078) | 0.00254 (-0.0022 – -0.0073) |
| **Modifiable NH Characteristics** | | | | |
| CNA to Nurse ratio |  |  |  |  |
| Middle Tertile | -0.0044 (-0.015 – 0.0064) | 0.000060 (-0.0048 – 0.0049) | -0.0092 (-0.021 – 0.0024) | -0.0024 (-0.0077 – 0.0030) |
| Highest Tertile | -0.0014 (-0.015 – 0.012) | 0.00049 (-0.0057 – 0.0067) | -0.0074 (-0.022 – 0.0072) | -0.0021 (-0.0089 – 0.0047) |
| RN to total nurse ratio |  |  |  |  |
| Middle Tertile | 0.007 (-0.0043 – 0.018) | -0.0023 (-0.0075 – 0.0029) | -0.0038 (-0.016 – 0.0083) | -0.0075* (-0.013 – -0.0019) |
| Highest Tertile | 0.03045* (0.015 – 0.046) | 0.00041 (-0.0068 – 0.0076) | 0.02196* (0.0050 – 0.039) | -0.003 (-0.011 – 0.0048) |
| Total nursing hours per resident day |  |  |  |  |
| Middle Tertile | 0.01796 (0.0038 – 0.032) | -0.0015 (-0.0077 – 0.0047) | -0.008 (-0.023 – 0.0070) | -0.001 (-0.0079 – 0.0059) |
| Highest Tertile | 0.00947 (-0.010 – 0.029) | -0.0054 (-0.014 – 0.0032) | -0.0132 (-0.034 – 0.0078) | 0.00311 (-0.0064 – 0.013) |
| Hospitalizations, per resident, per year |  |  |  |  |
| Middle Tertile | -0.0055 (-0.016 – 0.0052) | 0.00107 (-0.0038 – 0.0059) | -0.0034 (-0.015 – 0.0079) | -0.0002 (-0.0054 – 0.0050) |
| Highest Tertile | -0.0159 (-0.028 – 0.0042) | 0.00036 (-0.0050 – 0.0057) | -0.0135* (-0.026 – -0.0010) | -0.0008 (-0.0066 – 0.0050) |
| CNA hours per resident day |  |  |  |  |
| Middle Tertile | 0.00667 (-0.0072 – 0.021) | 0.00191 (-0.0042 – 0.0080) | 0.01549* (0.00077 – 0.0030) | 0.00538 (-0.0014 – 0.012) |
| Highest Tertile | -0.0009 (-0.021 – 0.019) | -0.0008 (-0.0093 – 0.0078) | 0.01816 (-0.0030 – 0.039) | -0.0015 (-0.011 – 0.0080) |
| LPN hours per resident day |  |  |  |  |
| Middle Tertile | 0.00363 (-0.0091 – 0.016) | -0.0006 (-0.0065 – 0.0052) | 0.01261 (-0.0011 – 0.026) | -0.0019 (-0.0083 – 0.0044) |
| Highest Tertile | 0.00181 (-0.016 – 0.019) | 0.00255 (-0.0053 – 0.010) | 0.00267 (-0.016 – 0.021) | -0.0032 (-0.012 – 0.0055) |
| Total physician extender FTEs/100 beds |  |  |  |  |
| Middle Tertile | -0.0056 (-0.016 – 0.0047) | -0.0026 (-0.0073 – 0.0020) | -0.0048 (-0.016 – 0.0062) | 0.00077 (-0.0043 – 0.0059) |
| Highest Tertile | -0.004 (-0.015 – 0.0069) | 0.00116 (-0.0038 – 0.0061) | -0.0101 (-0.022 – 0.0015) | -0.0024 (-0.0079 – 0.0031) |
| SLP total staff FTE |  |  |  |  |
| Middle Tertile | 0.01656* (0.0066 – 0.027) | 0.00473* (0.00023 – 0.0092) | 0.01016 (-0.00048 – 0.021) | 0.00717* (0.0023 – 0.012) |
| Highest Tertile | 0.01671* (0.0058 – 0.028) | 0.00697* (0.0020 – 0.012) | 0.01448* (0.0027 – 0.026) | 0.00978* (0.0043 – 0.015) |
| Social worker on staff/100 beds |  |  |  |  |
| Middle Tertile | -0.011 (-0.021 – 0.00077) | -0.0041 (-0.0088 – 0.00061) | -0.0122* (-0.023 – -0.0012) | -0.0056* (-0.011 – -0.00045) |
| Highest Tertile | 0.0093 (-0.0016 – 0.020) | 0.0083* (0.0033 – 0.013) | 0.00985 (-0.0019 – 0.022) | 0.00438 (-0.0010 – 0.0098) |
| Pressure ulcers |  |  |  |  |
| Middle Tertile | 0.0051 (-0.0051 – 0.015) | -0.0047* (-0.0093 – -0.00014) | -0.0021 (-0.013 – 0.0087) | -0.0041 (-0.0091 – 0.00089) |
| Highest Tertile | -0.0028 (-0.014 – 0.0082) | -0.0069* (-0.012 – -0.0020) | -0.0143* (-0.026 – -0.0026) | -0.0064* (-0.012 – -0.00098) |
| Restraints |  |  |  |  |
| Middle Tertile | -0.0047 (-0.015 – 0.0053) | -0.0005 (-0.0051 – 0.0042) | -0.0023 (-0.013 – 0.0084) | 0.00135 (-0.0037 – 0.0064) |
| Highest Tertile | -0.0067 (-0.017 – 0.0036) | -0.0035 (-0.0081 – 0.0012) | -0.0074 (-0.018 – 0.0035) | -0.0013 (-0.0064 – 0.0038) |

**Abbreviations**: NH, nursing home; CNA, certified nursing assistant; RN, registered nurse; LPN, licensed practical nurse; FTE, full-time equivalent; ADL, activities of daily living.

**Supplementary Table S3.** **County-Level Risk-Standardized Vaccination Rates of Long-Term Care Facility Residents Stability Analysis, by Season and Cohort**

|  | **2013-2014 Season** | | **2014-2015 Season** | |
| --- | --- | --- | --- | --- |
|  | **Short-Stay**  **Median (IQR)** | **Long-Stay**  **Median (IQR)** | **Short-Stay**  **Median (IQR)** | **Long-Stay**  **Median (IQR)** |
| % Vaccinated, original | 69.6 (62.8, 74.5) | 84.0 (80.8, 86.4) | 69.1 (62.0, 74.1) | 83.1 (79.7, 85.7) |
| % Vaccinated, >5 residents/county | 69.6 (62.5, 74.9) | 84.0 (80.8, 86.4) | 69.3 (61.7, 74.7) | 83.1 (79.7, 85.7) |
| % Vaccinated, >7 residents/county | 69.7 (62.4, 75.0) | 84.0 (80.8, 86.4) | 69.3 (61.7, 74.8) | 83.1 (79.7, 85.7) |
